# Multi-species co-circulation of adenoviruses identified by next generation sequencing during an outbreak in coastal Kenya in 2023

**DOI:** 10.1101/2024.03.21.24304701

**Authors:** Arnold W. Lambisia, Martin Mutunga, Esther N. Katama, Charles N. Agoti, Charlotte J. Houldcroft

**Affiliations:** KEMRI-Wellcome Trust Research Programme, Epidemiology and Demography Department, Kilifi, Kenya; Pwani University, School of Health and Human Sciences, Kilifi, Kenya; University of Cambridge, Department of Genetics, Downing Street, Cambridge, CB2 3EH, United Kingdom

**Author notes:** Corresponding author: Contact details: P.O. Box 230, Kilifi-80108, Kenya.

**Keywords:** Human adenovirus, ONT, enteric, Kenya, RespiCov

## Abstract

**Background:** Although seven human adenovirus (HAdV) species are known to exist,only F (types 40 and 41) and G, are identified as diarrhoeal disease agents. The role of other HAdV species in diarrhoeal disease remains unclear and data of their prevalence is limited. We describe HAdV species and types in hospitalised children with diarrhoea in coastal Kenya.

**Methods:** 329 stool samples collected between June 2022 and August 2023 from children aged <13-years were screened for HAdV using quantitative polymerase chain reaction (qPCR). Positive HAdV cases were genotyped by adenovirus primers from the RespiCoV panel by amplification, next generation sequencing followed by phylogenetic analysis.

**Results:** 65 samples (20%) tested HadV positive from which five HAdV species were identified. Other than HAdV F, other species included A, B, C and D; these were detected as either mono-detections or coinfections. Six HAdV F identified by NGS had been missed by our q PCR typing method. This appeared to be as a result of a 133-nucleotide deletion in the long fiber protein which abrogated a primer and probe binding site. Based on VESIKARI scores grading of diarrheal disease severity, 93% of the HAdV cases presented with severe disease. One child with an HAdV F infection died.

**Conclusion:** Our study shows the enormous diversity and clinical characteristics of HAdV species in children with diarrhoea in coastal Kenya. These data offers an opportunity to improve current diagnostic assays, increase knowledge of HAdV in Africa for control of outbreaks in the future.

## Background

Human adenoviruses (HAdV) are non-enveloped, double-stranded DNA viruses that belong to the *mastadenovirus* genus ^1^. To date, seven different species, A to G, and 114 different types of HAdV have been described by the human adenovirus working group (http://hadvwg.gmu.edu/). These HAdVs are associated with a variety of diseases presentation including gastroenteritis (F and G) ^2,3^, respiratory tract infections (A, B, C and E)^4^ and keratoconjunctivitis (D) ^5^.

HAdV types F40 and F41 are a common aetiology of mild to severe diarrhoea among children under the age of five years ^3,6–11^. However, other HAdV species such as A, B, C, D and E have been detected among diarrhoea cases although their contribution to diarrhoea disease is unclear ^3,7,10^. In China, HAdV B3 has been associated with diarrhoea (adjusted odds ratio = 9.205, *p* < 0.001) ^3^. In Canada, HAdV F40/41, detections had the highest attributable fraction (96%; 95% confidence interval (C.I), 92.3 to 97.7%) to diarrhoea symptoms compared to species A, B, C and E, but HAdV C1, C2, C5 and C6 were also attributed to 52% (95% C.I, 12 to 73%) of the symptoms ^10^. A previous study in Kenya reported predominance of HAdV species D and F in urban and rural settings respectively, among cases with diarrhoea; but other types including B3, B21, C2, C5 and C6 were also detected among diarrhoeal cases ^12^.

The KEMRI-Wellcome Trust Research Programme (KWTRP) has been conducting a prospective hospital-based rotavirus surveillance study at Kilifi County Hospital paediatric ward in coastal, Kenya ^6,13^. The prevalence of adenoviruses of any species among paediatric diarrhoea cases in coastal Kenya has been reported to be 15.9% (95% C.I 12.8 to 19.5). However, HAdV F only accounts for approximately half of the HAdV detections (7.3%, 95% C.I 5.2-10.1) with the rest of the HAdVs untyped ^13^.

This study aimed to genotype HAdV positive samples detected between June 2022 and August 2023 to determine the circulating non-F HAdVs and any HAdV-Fs that may have been missed by real-time PCR screening. HAdV genotyping is typically done by amplifying a region of the hexon gene, followed by Sanger sequencing ^14^. Here we used adenovirus primers from the RespiCoV panel ^15^, applying these primers to stool-derived nucleic acid extracts for the first time, and sequencing the amplicons on the Oxford Nanopore Technologies (ONT) platform.

## Methods

### Study site and population

The target population was children below the age of 13 years admitted to Kilifi County Hospital (KCH) who presented with diarrhoea as one of their illness symptoms i.e three or more loose stools in a 24-hour period ^16^.

### Laboratory Methods

#### Total Nucleic Acid (TNA) Extraction and Screening

TNA was extracted from 0.2 g (or 200 μl if liquid) of stool samples using QIAamp® Fast DNA Stool Mini kit (Qiagen, UK) as previously described. Pan-HAdV (forward primer: 5’-GCCCCAGTGGTCTTACATGCACATC -3’; probe: ‘FAM-TCGGAGTACCTGAGCCCGGGTCTGGTGCA-MGBNFQ’; and Reverse primer: 5’-GCCACGGTGGGGTTTCTAAACTT-3’) and HAdV-F (forward primer: 5’-CACTTAATGCTGACACGGGC-3’; probe: ‘FAM-TGCACCTCTTGGACTAGT-MGBNFQ’; and Reverse primer: 5’-ACTGGATAGAGCTAGCGGGC-3’) primers and probes, and TaqMan Fast Virus 1-Step Master Mix were used for screening as previously described ^17^. The thermocycling conditions were 95°C for 20 seconds and 35 cycles of 94°C for 15 seconds and 60°C for 30 seconds.

### DNA amplification

The primers used in amplification were adopted from the RespiCov panel ^15^. Briefly, 14 adenovirus primers were pooled into one tube and resuspended in nuclease free water to generate a 10μM working concentration. TNA from HAdV positive samples were amplifed using the Q5® Hot Start HighFidelity 2X Master Mix (NEB) kit. The master mix was prepared as follows: Q5® Hot Start High-Fidelity 2X Master Mix (6.25 μl), H2O (3 μl), HAdV Primer pool (2 μl), and DNA (1.25 μl). The reaction was then incubated on a thermocycler using the following conditions: 98°C for 30 seconds followed by 35 cycles of 98°C for 15 seconds, 65°C for 30 seconds and 72°C for 20 seconds and a final extension of 72°C for 5 minutes.

### Library Preparation and Oxford Nanopore Technologies (ONT) Sequencing

Library preparation was performed using the SQK-LSK114 ligation kit with SQK-NBD114.96 barcoding kit. Briefly, the amplicons were end-repaired, barcoded, and pooled into one tube, and adapters ligated to the library and the final library sequenced using the FLOW-MIN106D R9.4.1 flow cell on the GridION platform (ONT) for one hour.

### Long Fiber amplification and Illumina sequencing

Primers that could amplify the long fiber protein were obtained by picking forward (HAdV-F41_1kb_jh_85_LEFT:ACACTACAMTCCCCTTGACATCC) and reverse primers (HAdV-F41_1kb_jh_87_RIGHT:AAGAAAATGAGCAGCAGGGGATG) from the whole genome sequencing HAdV primers designed elsewhere ([Quick F41 WGS primers](https://github.com/quick-lab/HAdV/blob/main/HAdV-F41/v1.1/HAdV-F41_1kb_jh.primer.bed)). The mastermix reaction was prepared as described in the above section and incubated on a thermocycler using the following conditions: 98°C for 30 seconds followed by 35 cycles of 98°C for 15 seconds and 65°C for 5 minutes. The amplicons generated were used for library preparation using an Illumina library preparation kit as recommended by the manufacturer. Briefly, the amplicons were tagmented, indexed, and amplified. The libraries were then normalized, pooled, and sequenced as paired-end reads (2*150 bp).

### Data analysis Genotyping

The FASTQ reads from the GridION were trimmed using porechop v.0.2.4 and mapped to the HAdV reference genomes (DQ923122.2, NC_001460.1, NC_001454.1, NC_001405.1, AC_000006.1, AC_000018.1, AC_000008.1, NC_012959.1) using minimap2 v.2.24-r1122 (https://github.com/lh3/minimap2). Variant calling and consensus sequence generation was done using ivar v.1.3.1 (https://github.com/andersen-lab/ivar) with a minimum read depth of 20. Taxonomic classification was done using BLASTN (https://blast.ncbi.nlm.nih.gov/).

The generated consensus genomes were aligned with contemporaneous global HAdV sequences on GenBank using MAFFT (https://mafft.cbrc.jp/alignment/software/) and maximum likelihood trees generated using iqtree 2 (http://www.iqtree.org/). The trees were then annotated and visualized using ggtree (https://guangchuangyu.github.io/software/ggtree).

### Long fiber primer check validation

Short-read paired fastq data obtained from the Illumina MiSeq platform was trimmed using fastp with a phred scrore of q30. The cleaned reads were then mapped to the HAdV-F reference genome using bwa (https://github.com/lh3/bwa). The primers were then trimmed from the BAM files and consensus genomes generated using ivar.

For quality check, the cleaned reads were also assembled using a denovo approach using MetaSPAdes v3.13 (http://cab.spbu.ru/software/spades). The generated contigs were compared with consensus genomes from the reference guide approach.

Real-time PCR primer and probe sequences were then aligned to the generated consensus long fiber sequences to check for differences in binding sites using Geneious Prime® 2023.2.1 (https://www.geneious.com).

### Disease severity

The Vesikari Clinical Severity Scoring System Manual was used to estimate disease severity as previously described ^18^. The following parameters were used: maximum number of stools and vomiting per day, duration of diarrhoea and vomiting episodes in days, temperature, dehydration status and treatment. The Vesikari grading categories were mild, moderate, and severe for scores of <7, 7-10 and ≥ 11 respectively.

## Results

### HAdV Epidemiology

Between 3^rd^ June 2022 and 28^th^ August 2023, 329 children with diarrhoea as one of their illness symptoms were consented and gave stool samples for enteric viruses screening. A total of 65 (20%) cases had a HAdV detection in their sample when using a pan-adenovirus real-time PCR assay. Forty-three samples were successfully sequenced and genotyped using adenovirus RespiCoV primers to determine the circulating HAdV species and types in stool.

Single or multiple HAdV types were detected in the samples (**Figure 1**). Single HAdV type detections were as follows: F (n=13), C (n=12), D (n=3), B (n=2) and A (n=2). Codetection of HAdV samples was also observed in 10 samples: C and F (n=4), A and C (n=3), A and D (n=1), F and D (n=1), and C, F and D (n=1) (**Figure 1B**).

**Figure 1:**
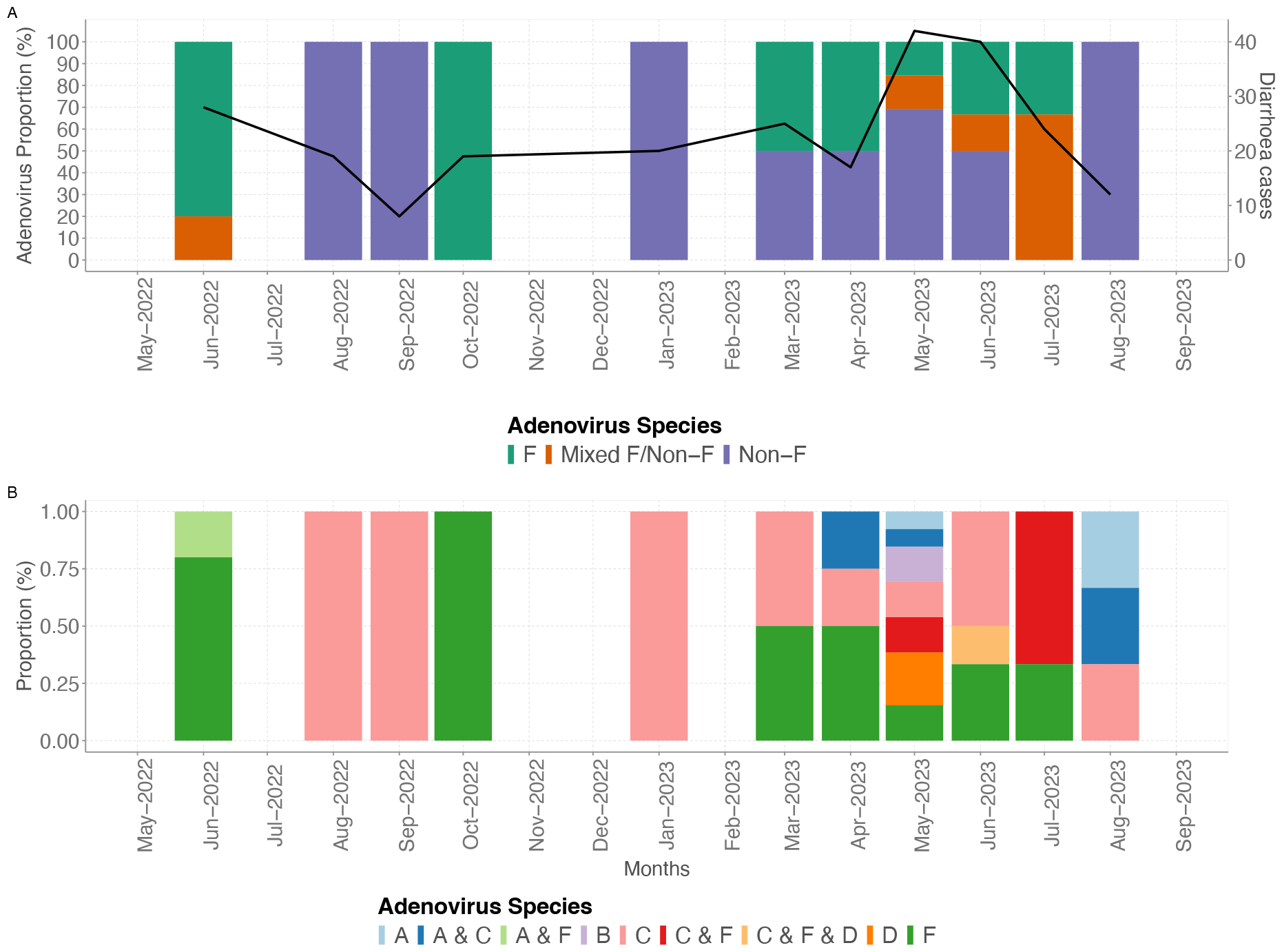
Temporal distribution of human adenoviruses in coastal Kenya between June 2022 and August 2023. A) Temporal plot showing distribution of F, mixed F (F and non-F coinfections) and non-F. The primary y axis shows the proportions of adenovirus species while the secondary y axis shows the total number of diarrhoea monthly cases. B) Temporal plot showing the distribution of HAdV species between June 2022 and August 2023.

The HAdV species were further characterized into types (**Figure 2**). Both HAdV F40 (n=7) and F41 (n=13) were detected in HAdV species F-positive cases. Within HAdV species A, A31 (n=4), A18 (n=2) and A12 (n=1) were identified. Only HAdV B7 was identified in species B. HAdV type C1 (n=5), C2 (n=5), C5 (n=5) and C89 (n=5) were identified within species HAdV C. Within HAdV D, D54 was identified in two samples while the rest of the sequences could not be genotyped further phylogenetically (**Figure 2**).

**Figure 2:**
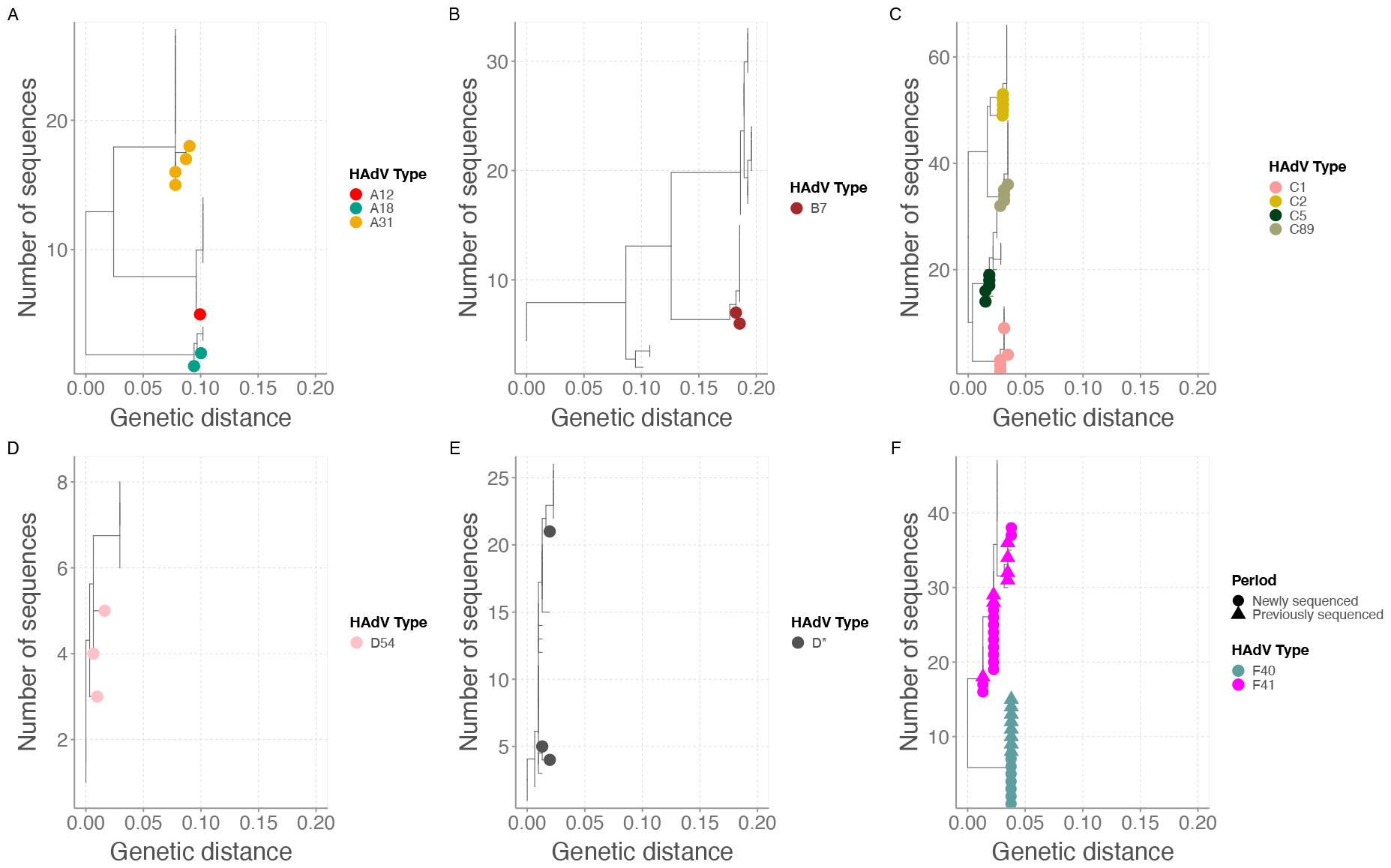
Maximum likelihood trees showing the divergence of detected HAdV types A) HAdV A B) HAdV B C) HAdV C D) HAdV D54 E) HAdV D* and F) HAdV F.

### Adenovirus outbreak in May 2023

In May 2023, there was a noticeable increase in adenovirus detections among children presenting with diarrhoea over the study period. From the 13 HAdV cases in May 2023, 69% of the cases were genotyped as non-HAdV F (D (n=3), B (n=2), C (n=2), A and C (n=1) and A (n=1)) and only 4 samples were genotyped as HAdV F. Notably, 38% of the 13 HAdV cases had coinfections with norovirus GII (n=4) and astrovirus (n=1).

### Demographic and clinical characteristics of HAdV cases

The majority of the HAdV cases were male (60.5%) and between the age of 12 to 59 months (65.1%). Coinfections with rotavirus A, norovirus GII, sapovirus and astrovirus were identified across the different HAdV species. Majority of the cases (93%) presented with severe disease, including all the non-F cases (**Table 1**). Only one fatality was identified in the HAdV F cases and none in the non-F cases (**Table 1**).

**Table 1:** Demographic and clinical characteristics of HAdV genotyped cases in coastal Kenya.

|  | HAdV F<br>(n=13) | HAdV Codetection F and non-F (n=7) | HAdV non-F<br>(n=23) | Total (n=43) |
| --- | --- | --- | --- | --- |
| **Sex** |  |  |  |  |
| Female | 7 (53.8%) | 2 (28.6%) | 6 (26.1%) | 15 (34.9%) |
| Male | 6 (46.2%) | 4 (57.1%) | 16 (69.6%) | 26 (60.5%) |
| No data | 0 (0.0%) | 1 (14.3%) | 1 (4.3%) | 2 (4.7%) |
| **Age (Months)** |  |  |  |  |
| **Median (interquartile range)** | 13 (7 – 21) | 11.5 (11-28.5) | 15.5 (10.2-20) | 14 (10-21) |
| **Age group (months)** |  |  |  |  |
| <12 | 1 (7.7%) | 2 (28.6%) | 3 (13.0%) | 6 (14.0%) |
| 12 - 23 | 5 (38.5%) | 1 (14.3%) | 9 (39.1%) | 15 (34.9%) |
| 24 - 59 | 5 (38.5%) | 2 (28.6%) | 6 (26.1%) | 13 (30.2%) |
| ≥60 | 2 (15.4%) | 2 (28.6%) | 5 (21.7%) | 9 (20.9%) |
| **Coinfections** |  |  |  |  |
| Rotavirus A | 1 (7.7%) | 2 (28.6%) | 3 (13.0%) | 6 (14.0%) |
| Norovirus GII | 0 (0.0%) | 1 (14.3%) | 4 (17.4%) | 5 (11.6%) |
| Sapovirus | 1 (7.7%) | 0 (0.0%) | 2 (8.7%) | 3 (7.0%) |
| Astrovirus | 0 (0.0%) | 1 (14.3%) | 0 (0.0%) | 1 (2.3%) |
| **Disease severity** |  |  |  |  |
| Moderate | 2 (15.4%) | 1 (14.3%) | 0 (0.0%) | 3 (7.0%) |
| Severe | 11 (84.6%) | 6 (85.7%) | 23 (100.0%) | 40 (93.0%) |
| **Outcome** |  |  |  |  |
| Alive | 12 (92.3%) | 6 (85.7%) | 22 (95.7%) | 40 (93.0%) |
| Dead | 1 (7.7%) | 0 (0.0%) | 0 (0.0%) | 1 (2.3%) |
| No data | 0 (0.0%) | 1 (14.3%) | 1 (4.3%) | 2 (4.7%) |

### Failure of HAdV-F real-time PCR Assay

The genotyping results detected six additional HAdV-F positives that were missed by real-time PCR. Successful sequencing of the long fiber protein where the real-time PCR primers bind showed that the sequences had a 133 base deletion that spanned the forward primer and probe binding regions leading to failure in the HAdV-F real-time PCR assay (**Figure 3**).

**Figure 3:**
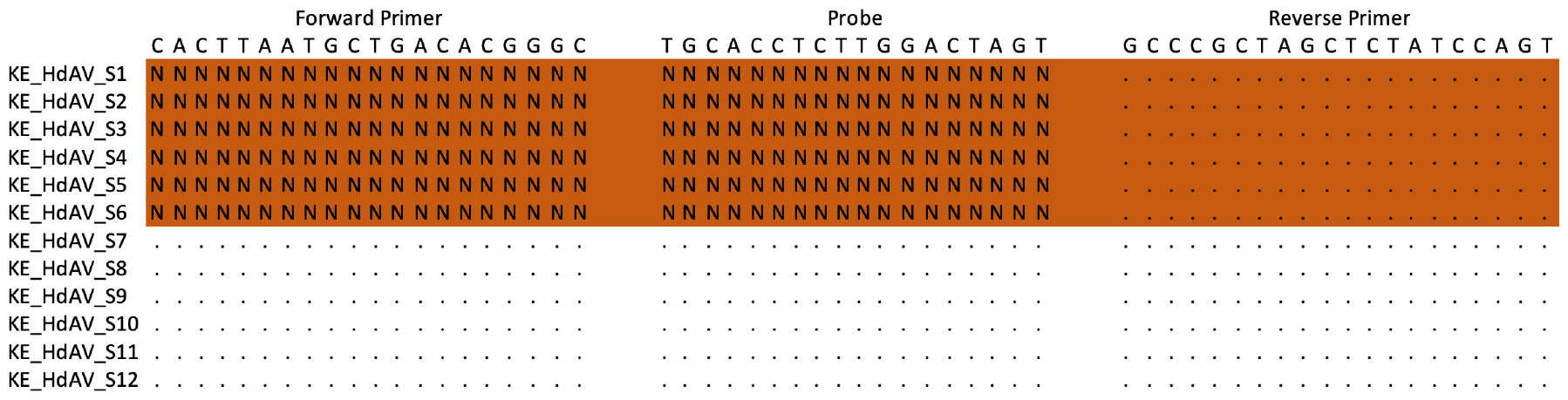
An alignment of HAdV F sequences from missed and detected HAdV F samples mapped to the primers and probe sequences. Dots show consensus and Ns show gaps in the primer and probe binding sites.

### Ct Value distribution

HAdV F cases had significantly lower Ct values compared to non-F cases (*p = 0*.*002*) (**Figure 4**). There was no significant difference in Ct value among HAdV F cases and cases that had codetections of HAdV-F and non-F. Eight non-F cases had a Ct value of less than 25 (high viral load) but six of these samples also had coinfections with norovirus GII, rotavirus A and sapovirus.

**Figure 4:**
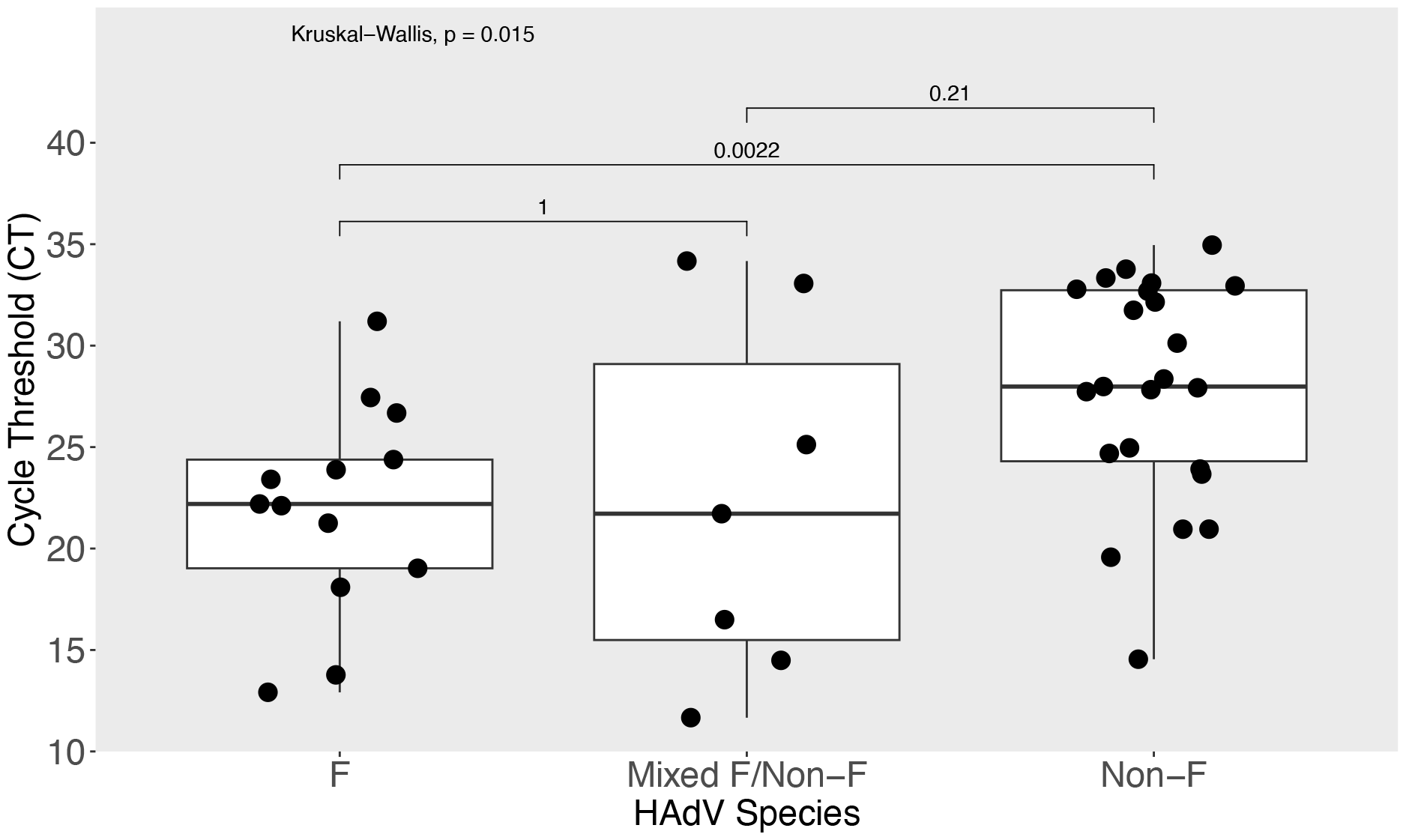
Comparison of HAdV viral load (inverse of cycle threshold value) among F, mixed F (coinfection of F and non-F) and non-F cases.

## Discussion

The study findings show that multiple HAdV species and types were in circulation between June 2022 and August 2023 in coastal Kenya and that our current real-time PCR primers for detecting HAdV F may be missing positive cases due to a 133-nucleotide deletion in the long fiber protein which abolishes a primer and probe binding site in some circulating variants. This is the first application of the adenovirus part of the RespiCov method to stool samples for genotyping enteric viral infections. Previous studies from China, Tunisia, Kenya and Brazil have reported detection of non-F HAdVs similar to our findings ^3,7,9,10,12,19,20^. Non-F types such as B3, C1, C2, C5 and C6 have been associated with increased risk of diarrhoea but direct causation is not yet clear ^3,10^. In this study, we did not detect HAdV B3 but detected the HAdV types C1 and C5; although we cannot conclude that they were the main cause of diarrhoea in these individuals. The RespiCov method used could not clearly genotype all HAdV species D phylogenetically. This is likely due to the relatively short ∼300 base pair hexon region that is amplified and the highly recombinatorial nature of species D HAdVs within the hexon locus ^21^.

Within HAdV F, there is a high divergence in the strains that have been reported to be circulating in Kenya and other regions across the globe ^17,22–24^. If this divergence occurs in primer and probe binding sites, nucleic acid amplification methods used for detection may be affected. The genotyping results in this study showed six HAdV F cases were missed by the F-specific real-time PCR assay. Amplification of the long fiber protein revealed a 133 base deletion that impacted the HAdV-F real-time PCR assay, and it is advisable that similar primers used in previous works ^13,17^ may be missing some positives and need redesigning, a phenomenon previously seen in SARS-CoV-2 ^25^, HIV ^26^ and *Chlamydia trachomatis* ^27^.

HAdV F samples had a significantly higher viral load (lower Ct value) compared to non-F samples, similar to previous studies ^10,19^. HAdV species F is highly associated with diarrhoea when the Ct value is below 22.7 and cases and controls can be discriminated at Ct 30.5 suggesting that the HAdV F detected in this study may be clearly associated with diarrhoea ^9,28^. Interestingly, the HAdV-F detections with Ct value above 30 had a rotavirus A coinfection suggesting that they could be the secondary cause of diarrhoea. The non-F HAdVs detected had a lower viral load and were detected with other enteric viruses including rotavirus A, norovirus GII and sapovirus suggesting that they may be associated with carriage due to prolonged shedding or gut contamination from respiratory infections rather than diarrhoea^29,30^.

This study had some limitations. First, there was no clinical data on the respiratory symptoms of the patients to help interpret the non-enteric HAdVs detected in stool samples which are usually associated with respiratory illnesses. Secondly, only four common enteric viruses were screened for, and these were frequently detected as coinfections in non-F cases. Screening for additional causes such as bacteria, parasites and helminths may have helped elucidate whether other enteric pathogens contribute to the detection of non-F cases.

In conclusion, there is a high diversity of HAdV types found among diarrhoea cases in coastal Kenya and this may reflect the situation in Africa where there is limited data. Timely and accurate genotyping of these HAdV cases is key for troubleshooting failure of molecular detection assays, estimation of diarrhoea prevalence associated with HAdV-F and implementation of interventions to reduce the burden of HAdV associated diarrhoea.

## Declarations

### Ethics approval and consent to participate

The research protocol for the study was approved at Kenya Medical Research Institute (KEMRI), by the Scientific and Ethics Review Unit (SSC#2861) in Nairobi, Kenya.

## Acknowledgement

We are grateful to the study participants who provided samples and members of the pathogen epidemiology and omics group at KEMRI-Wellcome Trust Programme who did sample collection and laboratory processing. This manuscript was written with the permission of Director KEMRI CGMRC.

## Data availability

The datasets used and/or analyzed during the current study are available from the KWTRP Research repository via https://doi.org/10.7910/DVN/XCHBND. The HAdV sequences were deposited on GenBank and can be accessed using the accession numbers PP318651-PP318703.

## Authors’ contributions

CAN and CJH sourced the study funding. CNA, CJH and AWL designed the study laboratory assay. AWL and MM did the laboratory experiments. EK and AWL managed the study data and did the data analysis. AWL, CAN and CJH wrote the first manuscript draft. All authors read, revised, and approved the final manuscript.

## References

1. Hulo C, de Castro E, Masson P, Bougueleret L, Bairoch A, Xenarios I, et al. ViralZone: a knowledge resource to understand virus diversity. Nucleic Acids Res [Internet]. 2011 Jan 1 [cited 2023 Dec 19];39(suppl_1):D576–82. Available from: 10.1093/nar/gkq901

2. Bányai K, Martella V, Meleg E, Kisfali P, Peterfi Z, Benkö M, et al. Searching for HAdV-52, the putative gastroenteritis-associated human adenovirus serotype in Southern Hungary. New Microbiol. 2009;32(2):185–8.

3. Qiu F zhou, Shen X xin, Li G xia, Zhao L, Chen C, Duan S xia, et al. Adenovirus associated with acute diarrhea: A case-control study. BMC Infect Dis. 2018;18(1):1–7.

4. Lynch J, Kajon A. Adenovirus: Epidemiology, Global Spread of Novel Serotypes, and Advances in Treatment and Prevention. Semin Respir Crit Care Med [Internet]. 2016 Aug 3;37(04):586–602. Available from: 10.1055/s-0036-1584923

5. Adhikary AK, Banik U. Human adenovirus type 8: The major agent of epidemic keratoconjunctivitis (EKC). J Clin Virol [Internet]. 2014 Dec;61(4):477–86. Available from: https://linkinghub.elsevier.com/retrieve/pii/S1386653214004028

6. Lambisia AW, Murunga N, Mutunga M, Cheruiyot R, Maina G, Makori TO, et al. Temporal changes in the positivity rate of common enteric viruses among paediatric admissions in coastal Kenya, during the COVID-19 pandemic, 2019–2022. Gut Pathog [Internet]. 2024 Jan 4;16(1):2. Available from: https://gutpathogens.biomedcentral.com/articles/10.1186/s13099-023-00595-4

7. Afrad MH, Avzun T, Haque J, Haque W, Hossain ME, Rahman AFMR, et al. Detection of enteric-and non-enteric adenoviruses in gastroenteritis patients, Bangladesh, 2012-2015. J Med Virol. 2018;90(4):677–84.

8. Ghebremedhin B. Human adenovirus: Viral pathogen with increasing importance. Eur J Microbiol Immunol. 2014;4(1):26–33.

9. Huang Z, He Z, Wei Z, Wang W, Li Z, Xia X, et al. Correlation between prevalence of selected enteropathogens and diarrhea in children: A case-control study in China. Open Forum Infect Dis. 2021;8(10):1–7.

10. Pabbaraju K, Tellier R, Pang XL, Xie J, Lee BE, Chui L, et al. A Clinical Epidemiology and Molecular Attribution Evaluation of Adenoviruses in Pediatric Acute Gastroenteritis: a Case-Control Study on behalf of the Alberta Provincial Pediatric EnTeric Infection TEam (APPETITE). 2020;(December):1–11. Available from: 10.1128/JCM.02287

11. Binder AM, Biggs HM, Haynes AK, Chommanard C, Lu X, Dean ;, et al. Morbidity and Mortality Weekly Report Human Adenovirus Surveillance-United States, 2003-2016. MMWR Morb Mortal Wkly Rep. 2017;66(39):1039–1042.

12. Magwalivha M, Wolfaardt M, Kiulia NM, van Zyl WB, Mwenda JM, Taylor MB. High prevalence of species D human adenoviruses in fecal specimens from Urban Kenyan children with diarrhea. J Med Virol [Internet]. 2010 Jan;82(1):77–84. Available from: 10.1002/jmv.21673

13. Agoti CN, Curran MD, Murunga N, Ngari M, Muthumbi E, Lambisia AW, et al. Differences in epidemiology of enteropathogens in children pre-and post-rotavirus vaccine introduction in Kilifi, coastal Kenya. Gut Pathog [Internet]. 2022 Dec 1;14(1):32. Available from: 10.1186/s13099-022-00506-z

14. Lu X, Erdman DD. Molecular typing of human adenoviruses by PCR and sequencing of a partial region of the hexon gene. Arch Virol. 2006;151(8):1587–602.

15. Brinkmann A, Uddin S, Ulm SL, Pape K, Förster S, Enan K, et al. RespiCoV: Simultaneous identification of Severe Acute Respiratory Syndrome Coronavirus 2 (SARS-CoV-2) and 46 respiratory tract viruses and bacteria by amplicon-based Oxford-Nanopore MinION sequencing. PLoS One. 2022;17(3 March):1–11.

16. World Health organization - WHO. The Treatment of diarrhoea : a manual for physicians and other senior health workers. --4th rev [Internet]. Vol. 17. 2005 [cited 2023 Oct 2]. p. 562–3. Available from: https://www.who.int/publications/i/item/9241593180

17. Lambisia AW, Makori TO, Mutunga M, Cheruiyot R, Murunga N, Quick J, et al. Genomic epidemiology of human adenovirus F40 and F41 in coastal Kenya: A retrospective hospital-based surveillance study (2013-2022). Virus Evol. 2023;9(1):vead023.

18. Lewis K. VAD_vesikari_scoring_manual.pdf. 2011.

19. do Nascimento LG, Fialho AM, de Andrade J da SR, de Assis RMS, Fumian TM. Human enteric adenovirus F40/41 as a major cause of acute gastroenteritis in children in Brazil, 2018 to 2020. Sci Rep [Internet]. 2022;12(1):1–12. Available from: 10.1038/s41598-022-15413-1

20. Bouazizi A, Ben Hadj Fredj M, Bennour H, Jerbi A, Ouafa kallala, Fodha I, et al. Molecular analysis of adenovirus strains responsible for gastroenteritis in children, under five, in Tunisia. Heliyon [Internet]. 2024;10(1):e22969. Available from: 10.1016/j.heliyon.2023.e22969

21. Walsh MP, Chintakuntlawar A, Robinson CM, Madisch I, Harrach B, Hudson NR, et al. Evidence of Molecular Evolution Driven by Recombination Events Influencing Tropism in a Novel Human Adenovirus that Causes Epidemic Keratoconjunctivitis. Markotter W, editor. PLoS One [Internet]. 2009 Jun 3;4(6):e5635. Available from: https://dx.plos.org/10.1371/journal.pone.0005635

22. Gökng J, Cordes AK, Steinbrück L, Heim A. Molecular Phylogeny of human adenovirus type 41 lineages. bioRxiv [Internet]. 2022;2022.05.30.493978. Available from: 10.1101/2022.05.30.493978v1 %0A https://www.biorxiv.org/content/10.1101/2022.05.30.493978v1.abstract

23. Chandra P, Lo M, Mitra S, Banerjee A, Saha P, Okamoto K, et al. Genetic characterization and phylogenetic variations of human adenovirus-F strains circulating in eastern India during 2017–2020. J Med Virol [Internet]. 2021 Nov 29;93(11):6180–90. Available from: 10.1002/jmv.27136

24. Maes M, Khokhar F, Wilkinson SA, Smith A, Kovalenko G, Dougan G, et al. Enteric adenovirus F41 genetic diversity comparable to pre-COVID-19 era: validation of a multiplex amplicon-MinION sequencing method. OSFPREPRINTS [Internet]. 2022;Preprint:1–23. Available from: https://osf.io/6jku5

25. Volz E, Mishra S, Chand M, Barrett JC, Johnson R, Geidelberg L, et al. Assessing transmissibility of SARS-CoV-2 lineage B.1.1.7 in England. Nature [Internet]. 2021 May 13;593(7858):266–9. Available from: https://www.nature.com/articles/s41586-021-03470-x

26. Müller B, Nübling CM, Kress J, Roth WK, De Zolt S, Pichl L. How safe is safe: new human immunodeficiency virus Type 1 variants missed by nucleic acid testing. Transfusion [Internet]. 2013 Oct 17;53(10pt2):2422–30. Available from: 10.1111/trf.12298

27. Ripa T, Nilsson PA. A Chlamydia trachomatis Strain With a 377-bp Deletion in the Cryptic Plasmid Causing False-Negative Nucleic Acid Amplification Tests. Sex Transm Dis [Internet]. 2007 May;34(5):255–6. Available from: https://journals.lww.com/00007435-200705000-00001

28. Liu J, Plags-Mills JA, Juma J, Kabir F, Nkeze J, Okoi C, et al. Use of quantitative molecular diagnostic methods to identify causes of diarrhoea in children: a reanalysis of the GEMS case-control study. Lancet. 2016;

29. Bartholomeusz A, Locarnini S. Associated With Antiviral Therapy. Antivir Ther. 2006;55(November 2005):52–5.

30. Kim JS, Lee SK, Ko DH, Hyun J, Kim HS, Song W, et al. Associations of adenovirus genotypes in Korean acute gastroenteritis patients with respiratory symptoms and intussusception. Biomed Res Int. 2017;2017.

